# Sero-epidemiological evaluation of malaria transmission in The Gambia before and after mass drug administration

**DOI:** 10.1101/2020.04.09.20059774

**Authors:** Lindsey Wu, Julia Mwesigwa, Muna Affara, Mamadou Bah, Simon Correa, Tom Hall, James G. Beeson, Kevin K.A. Tetteh, Immo Kleinschmidt, Umberto D’Alessandro, Chris Drakeley

**Affiliations:** London School of Hygiene and Tropical Medicine (LSHTM), Faculty of Infectious Tropical Diseases, Department of Infection Biology. London WC1E 7HT, UK; Medical Research Council Unit The Gambia at London School of Hygiene and Tropical Medicine. Fajara, The Gambia; Bernhard Nocht Institute for Tropical Medicine (BNITM). Arusha, Tanzania; St. George’s University of London (SGUL). SW17 0RE, London, United Kingdom; Burnet Institute. Melbourne, Victoria, 3004, Australia; Central Clinical School, Monash University, Victoria, Australia; Department of Medicine, University of Melbourne, Victoria, Australia; London School of Hygiene and Tropical Medicine (LSHTM), Faculty of Epidemiology and Population Health, Department of Infectious Disease Epidemiology. London WC1E 7HT, UK; School of Pathology, Wits Institute for Malaria Research, Faculty of Health Science, University of Witwatersrand. Johannesburg, South Africa

**Keywords:** malaria, serology, mass drug administration, surveillance

## Abstract

**Background:** As The Gambia aims to achieve elimination by 2030, serological assays are a useful surveillance tool to monitor trends in malaria incidence and evaluate community-based interventions.

**Methods:** Within a mass drug administration (MDA) study in The Gambia, where reduced malaria infection and clinical disease were observed after the intervention, a serological sub-study was conducted in four study villages. Spatio-temporal variation in transmission was measured with a panel of recombinant *Pf* antigens on a multiplexed bead-based assay. Village-level antibody levels were quantified as under-15 sero-prevalence, sero-conversion rates, and age-adjusted antibody acquisition rates. Antibody levels prior to MDA were assessed for association with persistent malaria infection after community chemoprophylaxis.

**Results:** Seasonal changes in antibodies to Etramp5.Ag1 were observed in children under 15 in two transmission settings – the West Coast and Upper River Regions (4·32% and 31·30% *Pf* prevalence, respectively). At the end of the malaria season, short-lived antibody responses to Etramp5.Ag1, GEXP18, HSP40.Ag1, EBA175 RIII-V, and Rh2.2030 were lower amongst 1-15 year olds in the West Coast compared to the Upper River, reflecting known differences in transmission. Prior to MDA, individuals in the top 50^th^ percentile of antibody levels had two-fold higher odds of clinical malaria during the transmission season, consistent with previous findings where individuals infected pre-MDA had 2-fold higher odds of re-infection post-MDA.

**Conclusion:** Serological markers can serve dual functions as indicators of malaria exposure and incidence. Further studies, particularly cluster randomised trials, can help establish standardised serological protocols to measure transmission across endemic settings.

## Background

In malaria elimination settings, heterogeneity and hotspots of transmission are increasingly prevalent as disease burden declines.^1–5^ At low transmission, large proportions of the population can remain malaria free for years, while subpopulations experience multiple episodes^3,4,6^. This presents significant challenges for the implementation of malaria interventions and clinical trials designed to evaluate them^7,8^; if untargeted, residual transmission is likely to persist.^5,9–11^ Therefore, understanding why malaria has significantly changed in some settings, but remains unyielding in others, will be critical for guiding investments in malaria control in Sub-Saharan Africa.^12,13^

The Gambia has a long history of research on the heterogeneity of malaria in West Africa. Entomological and clinical data have shown spatial variation in malaria infections within two kilometres of mosquito breeding sites, where immune differences between households was hypothesised to drive differences in clinical outcomes.^14^ Studies in The Gambia have also demonstrated the impact of insecticide-treated bed nets on childhood mortality,^15,16^ with additional reductions in clinical malaria when combined with chemoprophylaxis.^17^ Subsequently, there have been rapid declines in incidence through the scale-up of control interventions - improved diagnosis and treatment, distribution of long-lasting insecticidal nets, indoor residual spraying, and seasonal malaria chemoprevention. However, micro-epidemiological variations in transmission remain, ^18,19^ which are increasingly relevant as The Gambia aims for malaria elimination by 2030. Sufficiently sensitive diagnostics will be critical to reach this target, helping to identify foci of transmission and target interventions.

Serological studies in The Gambia have described country-wide heterogeneities in malaria transmission, where schools surveys found strong correlations between sero-prevalence and microscopy-detectable parasitaemia.^20^ More recently, studies based on data from Uganda and Mali have identified several serological markers of *Plasmodium falciparum (Pf)* malaria exposure that are strongly associated with clinical incidence^21^. These measures of sero-incidence have the potential to supplement existing surveillance tools to monitor transmission and evaluate the effectiveness of community interventions.

While serology has been used in research settings to measure malaria transmission for some time, standardised antigen panels for routine surveillance have yet to be established. Furthermore, gaps in our understanding of how antibody levels reflect changes in transmission still remain. Therefore, this study evaluates the use of antibody responses as a measure of malaria exposure and incidence, with the aim to inform the future design of surveillance strategies. We used serological markers to measure spatio-temporal variation in transmission in a subset of four study villages from the Malaria Transmission Dynamics Study in The Gambia, which assessed epidemiological trends in six village pairs,^18,19^ followed by two years of mass drug administration (MDA) with dihydroartemisnin-piperaquine (DHA-PQ).^22^ A panel of recombinant *Pf* antigens was used to quantify antibody levels using a multiplexed immuno-assay. Village-level serological profiles were used to describe seasonal and geographical variation in antibody responses, and the association between antibody levels prior to MDA and persistent malaria infection during the transmission season after MDA was also estimated.

## Methods

### Data and sampling

A prospective cohort study was conducted from 2013 to 2015 in six village pairs across five administrative regions - West Coast (WCR), North Bank (NBR), Lower River (LRR), Central River (CRR) and Upper River (URR) Regions. *Pf* prevalence measured by PCR ranged from 2.27 to 19.60% in the Central River and Upper River Regions respectively. As previously described by Mwesigwa et al,^19^ the study aimed to understand the dynamics of malaria infection and the impact of annual MDA. All residents aged more than six months were enrolled. Monthly surveys were conducted throughout the transmission season (June to December), the dry season (April 2014) and before MDA implemented in May and June. In 2014 and 2015, one round of MDA was conducted, with DHA-PQ administered to individuals aged between six months and 75 years, according to weight-based dosing guidelines, over 6 to 14 days between May and June in the six pairs of study villages. Outcomes included incidence of clinical disease, prevalence of *Pf* infection measured by polymerase chain reaction (PCR), and factors associated with infection post-MDA.

Blood samples were collected by finger prick for haemoglobin measurement, blood smear for malaria diagnosis by microscopy, and molecular and serological analysis by dried blood spot (DBS) on filter paper (Whatman 3 Corporation, Florham Park, NJ, USA). Clinical malaria cases were identified by passive case detection (PCD) at local health facilities or in villages by study nurses; clinical malaria was defined as history of fever in the previous 24 hours or axillary temperature ≥ 37·5°C and a positive rapid diagnostic test (RDT) result (Paracheck *Pf*, Orchid Biomedical System, India).

Serological analysis included all available samples from the WCR in Besse and N’Demban from surveys done at the start of the transmission season in July 2013 (N=537) and at the end of the season in December 2013 (N=526). In the URR, analysis included all samples collected in Njaiyal and Madina Samako in July 2013 (N=779), December 2013 (N=632), April (dry season) 2014 (N=823), and December 2014 (N=757). These regions are at the extreme of transmission intensity found in The Gambia – low in WCR and moderate in URR - with months selected at the start and end of the transmission season. Samples from clinical PCD cases were linked by study participant identification code to samples from the same individuals collected during routine monthly surveys. Samples collected as part of the Malaria Transmission Dynamics Study and subset of samples for serological analysis are described in Figure 1.

**Figure 1.**
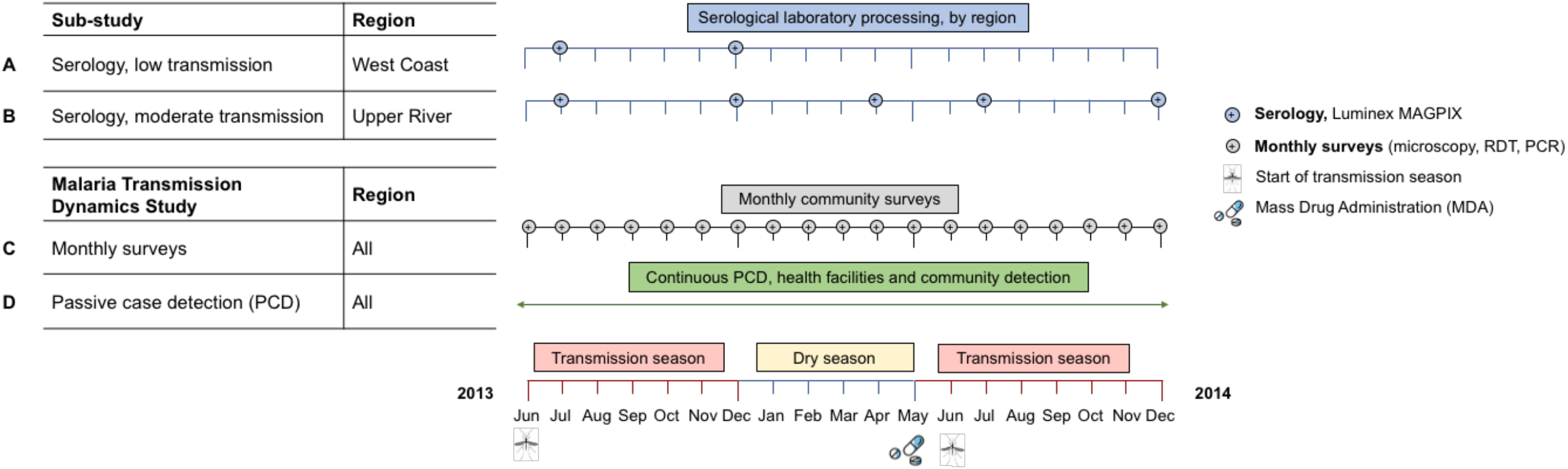
Study timelines. Malaria Transmission Dynamics Study timeline shown in black and green. Serological study timeline shown in blue for West Coast and Upper River Regions (low and moderate transmission settings, respectively). Samples for serological analysis were processed on the Luminex MAGPIX, samples from monthly surveys were analysed using microscopy, rapid diagnostic tests (RDTs) and polymerase chain reaction (PCR). As part of the Malaria Transmission Dynamics Study, one round of mass drug administration (MDA) was administered from May – June in both 2014 and 2015 (2015 not shown as no samples from this year were used for serological analysis). Transmission season is approximately June - December each year.

### Laboratory procedures

A full description of laboratory methods, including antigen amino acid range, selection and expression platform, are detailed in Wu et al^23^ and Mwesigwa et al^24^. Briefly, serum samples were eluted from 6mm DBS (8 µL whole blood equivalent) and prepared as a 1:500 dilution. Two wells on each plate containing only antigen-coupled beads and sample buffer were included to measure background signal. A pool of 22 serum samples from malaria hyper-immune individuals in The Gambia were used as positive controls in a 6-point 5-fold serial dilution (1:10 – 1:31,250) and plasma samples from European malaria-naive adults (1:500 dilution) as negative controls. Serum samples were processed on the Luminex MAGPIX, with mean fluorescence intensity (MFI) values between plates normalised according to the protocol described in Wu et al.^25^

For diagnostic PCR, DNA was extracted from three 6-mm DBS (4 µL whole blood equivalent) using the automated QIAxtractor robot (Qiagen). Negative and positive (3D7) controls were included to control for cross contamination and DNA extraction efficiency, respectively. Extracted DNA was used in a nested PCR, amplifying multi-copy *Plasmodium* ribosomal RNA gene sequences using genus and species-specific primers. All PCR products were run using the QIAxcel capillary electrophoresis system (Qiagen).

### Statistical analyses

Antibody responses were summarised at village level and used to describe geographical differences in transmission between WCR (low transmission) and URR (moderate-high transmission). Within each region, temporal changes were assessed using antibody responses at the start and end of transmission season (July and December 2013) and the dry season (April 2014), represented with the methods as summarised below.

#### Sero-prevalence and sero-conversion rate

For antigens associated with longer lived antibody responses - *P*.*falciparum* merozoite surface antigen 1 19-kDa carboxy-terminal region (*Pf*MSP1^19^) and *P*.*falciparum* apical membrane antigen 1 (*Pf*AMA1), the distribution of MFI for all serological samples was characterised through a two-component Gaussian mixture model to define distributions of negative and positive antibody levels. Sero-positivity thresholds were defined as the mean log MFI plus two standards deviations of the negative distribution.^26^ For antigens associated with shorter-lived antibodies – early transcribed membrane protein 5 (Etramp5.Ag1), gametocyte export protein 18 (GEXP18), heat shock protein 40 (HSP40.Ag1), erythrocyte binding antigen 175 RIII-V (EBA175), reticulocyte binding protein homologue 2 (Rh2.2030) - sero-positivity thresholds were defined by the mean log MFI plus three standard deviations of 71 malaria-naïve European blood donors used as negative controls.

Age-adjusted sero-conversion rates (SCRs), the annual rate at which sero-negative individuals become sero-positive, were estimated for antigens associated with long-lived antibody response (*Pf*MSP1^19^, *Pf*AMA1). A reversible catalytic model was fitted to age-adjusted sero-positivity data using maximum likelihood methods.^27^ A common sero-reversion rate for each antigen was assumed across all villages, which was estimated based on fitting a single sero-catalytic model to all individuals and fixing the sero-reversion rate in subsequent model fits. Additionally, sero-prevalence was estimated for children aged 1-15 years. Given the lower likelihood of long-lived antibodies from previous malaria exposure, sero-positivity in this age group is more likely to reflect recent infection.^20,28^

#### Age-dependent antibody acquisition

Antibody acquisition models^29,30^ were used to estimate age-dependent changes in antibody levels. Two variants of the model were used – one assuming a constant rate of increasing antibody levels with age and one assuming differing antibody acquisition rates between age groups (Supplementary Methods). Area Under the Antibody Acquisition Curve (AUC) values were calculated based on the model fit for each village and survey month, representing cumulative antibody levels across all ages.^31^ Within region AUCs were compared between the start and end of transmission season as well as between regions at each time point. Strength of evidence for observed differences are presented as p-values, derived from the posterior distributions of predicted antibody levels over a specified age range.

#### Dry season antibody levels and odds of infection post-MDA

Individual antibody levels in April 2014 prior to MDA were assessed by comparing observed and expected MFI value as predicted by the antibody acquisition fit for individuals in URR-South. For individuals with above average MFI values for their age group, the difference between observed and expected logMFI was calculated and classified to below or above median of residual MFI antibody response. Logistic regression was used to estimate the odds of clinical malaria (passively detected via the health facility or study nurses in the community) or asymptomatic *Pf* infection (actively detected using PCR from monthly survey samples) during the transmission season following MDA (October – December 2014) amongst individuals below and above median residual dry season antibody levels, compared to individuals at or below expected antibody responses, allowing for clustering at the household/compound. Multivariate analyses also adjusted for age, LLIN use, and MDA adherence using generalised linear mixed-effects models with the ‘glm’ and ‘glmer’ functions in the ‘lme4’ v1.1-21 package in R version 3.6.1.

## Results

### Sero-prevalence in children by geographical region and transmission season

Variation in sero-prevalence between geographical region and transmission season differed by antigen. More specifically, between July and December (start and end of the transmission season), seroprevalence to Etramp5.Ag1 in children aged 1-15 years in the West Coast Region increased by 4·6% (95%CI 1·0 - 8·3) and by 2·9% (95%CI 0·0 - 5·8) for *Pf*MSP1^19^ (Table 1, Figure 2, Supplementary Table 1). In the URR, there was statistically strong evidence for increases in under-15 sero-prevalence between July and December for all antigens (Supplementary Table 1). Mean sero-prevalence in the URR in December 2014, the transmission season after MDA, was lower for *Pf*MSP1^19^, *Pf*AMA1, Etramp5.Ag1, Rh2.2030 and EBA175, compared to December 2013 before implementation of MDA, but results were not statistically significant (Supplementary Table 1).

**Figure 2.**
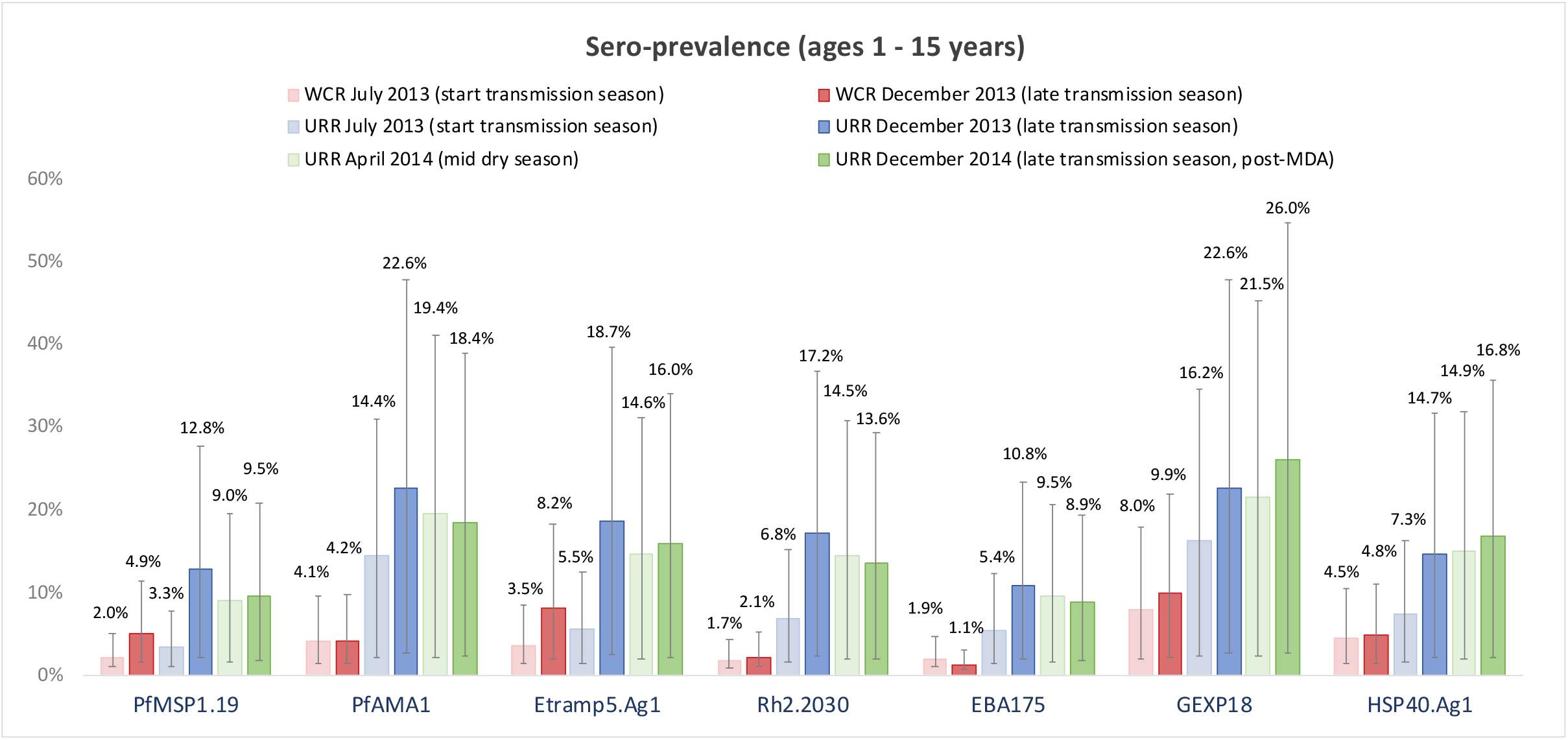
Sero-prevalence in children ages 1-15 years by antigen, geographical region, and transmission season. Mean sero-prevalence estimates are shown for the West Coast Region (WCR) for July 2013 (light red) and December 2013 (dark red), and the Upper River Region (URR) for July 2013 (light blue), December 2013 (dark blue), April 2014 (light green), and December 2014 (dark green). 95% confidence intervals are indicated by the vertical bars.

**Table 2.**
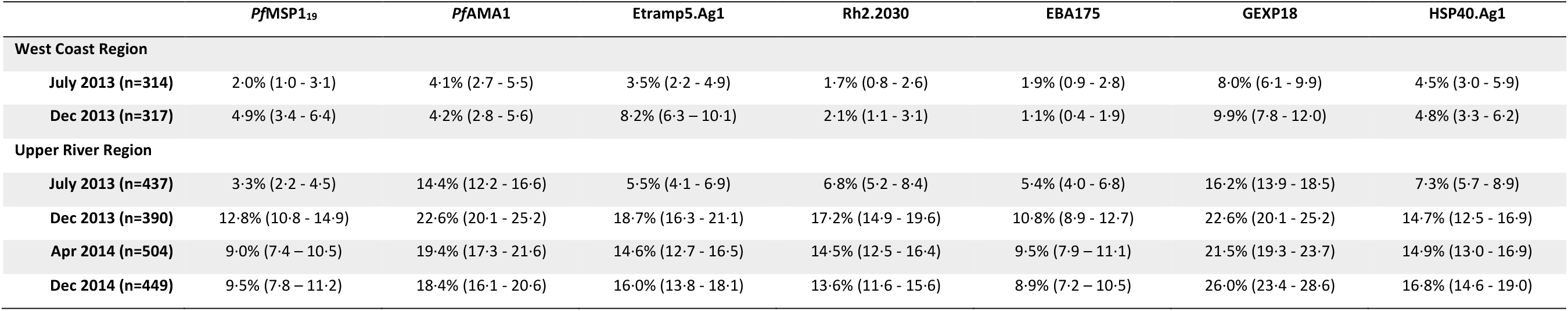
Sero-prevalence in children aged 1-15 years by antigen, geographical region, and transmission season. Mean sero-prevalence estimates are shown for the West Coast Region (WCR) for July 2013 and December 2013, and the Upper River Region (URR) for July 2013, December 2013, April 2014, and December 2014. Mean and 95% confidence intervals (parentheses) are shown as percentages.

|  | *Pf*MSP1<sub>19</sub> | *Pf*AMA1 | Etramp5.Ag1 | Rh2.2030 | EBA175 | GEXP18 | HSP40.Ag1 |
| --- | --- | --- | --- | --- | --- | --- | --- |
| **West Coast Region** |  |  |  |  |  |  |  |
| **July 2013 (n=314)** | 2.0% (1.0 - 3.1) | 4.1% (2.7 - 5.5) | 3.5% (2.2 - 4.9) | 1.7% (0.8 - 2.6) | 1.9% (0.9 - 2.8) | 8.0% (6.1 - 9.9) | 4.5% (3.0 - 5.9) |
| **Dec 2013 (n=317)** | 4.9% (3.4 - 6.4) | 4.2% (2.8 - 5.6) | 8.2% (6.3 - 10.1) | 2.1% (1.1 - 3.1) | 1.1% (0.4 - 1.9) | 9.9% (7.8 - 12.0) | 4.8% (3.3 - 6.2) |
| **Upper River Region** |  |  |  |  |  |  |  |
| **July 2013 (n=437)** | 3.3% (2.2 - 4.5) | 14.4% (12.2 - 16.6) | 5.5% (4.1 - 6.9) | 6.8% (5.2 - 8.4) | 5.4% (4.0 - 6.8) | 16.2% (13.9 - 18.5) | 7.3% (5.7 - 8.9) |
| **Dec 2013 (n=390)** | 12.8% (10.8 - 14.9) | 22.6% (20.1 - 25.2) | 18.7% (16.3 - 21.1) | 17.2% (14.9 - 19.6) | 10.8% (8.9 - 12.7) | 22.6% (20.1 - 25.2) | 14.7% (12.5 - 16.9) |
| **Apr 2014 (n=504)** | 9.0% (7.4 - 10.5) | 19.4% (17.3 - 21.6) | 14.6% (12.7 - 16.5) | 14.5% (12.5 - 16.4) | 9.5% (7.9 - 11.1) | 21.5% (19.3 - 23.7) | 14.9% (13.0 - 16.9) |
| **Dec 2014 (n=449)** | 9.5% (7.8 - 11.2) | 18.4% (16.1 - 20.6) | 16.0% (13.8 - 18.1) | 13.6% (11.6 - 15.6) | 8.9% (7.2 - 10.5) | 26.0% (23.4 - 28.6) | 16.8% (14.6 - 19.0) |

### Sero-conversion rate between geographical regions and transmission seasons

Differences in SCR between geographical region and transmission season varied by antigen (Figure 3, Supplementary Table 2). In WCR, changes in antibody responses between transmission seasons were larger for *Pf*MSP1^19^ (0·0147 increase in SCR between July and December) compared to *Pf*AMA1 (0·0007 increase in SCR). In URR, seasonal differences between July and December 2013 for both antigens were larger than the WCR, with an increase in *Pf*MSP1^19^ SCR of 0·0405 and increase of 0·0301 in *Pf*AMA1 SCR. There was no significant change in SCR between December 2013 and April 2014 for either antigen.

**Figure 3.**
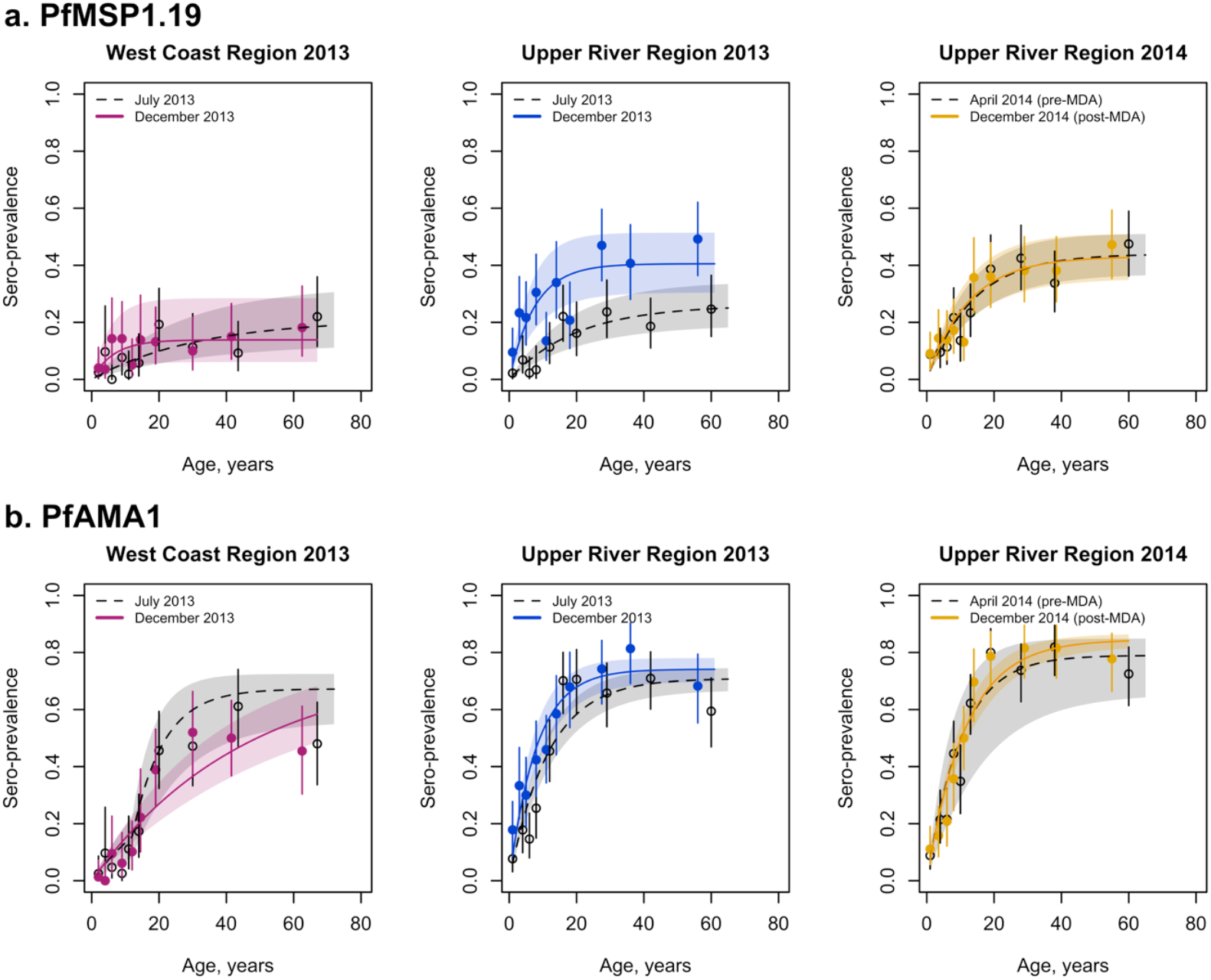
*Pf*MSP1.19 and *Pf*AMA1 sero-conversion rates by transmission season and geographical region. Mean and 95% confidence intervals of sero-prevalence for each age group are shown as circles and vertical lines, respectively. Mean fit of the reverse catalytic model is shown as a solid line for the end of the transmission season (December 2013 and 2014) and dotted line for the dry season (April 2014) or start of the transmission season (July 2013). Shaded regions are the 95% credible interval of the model fit.

**Figure 4.**
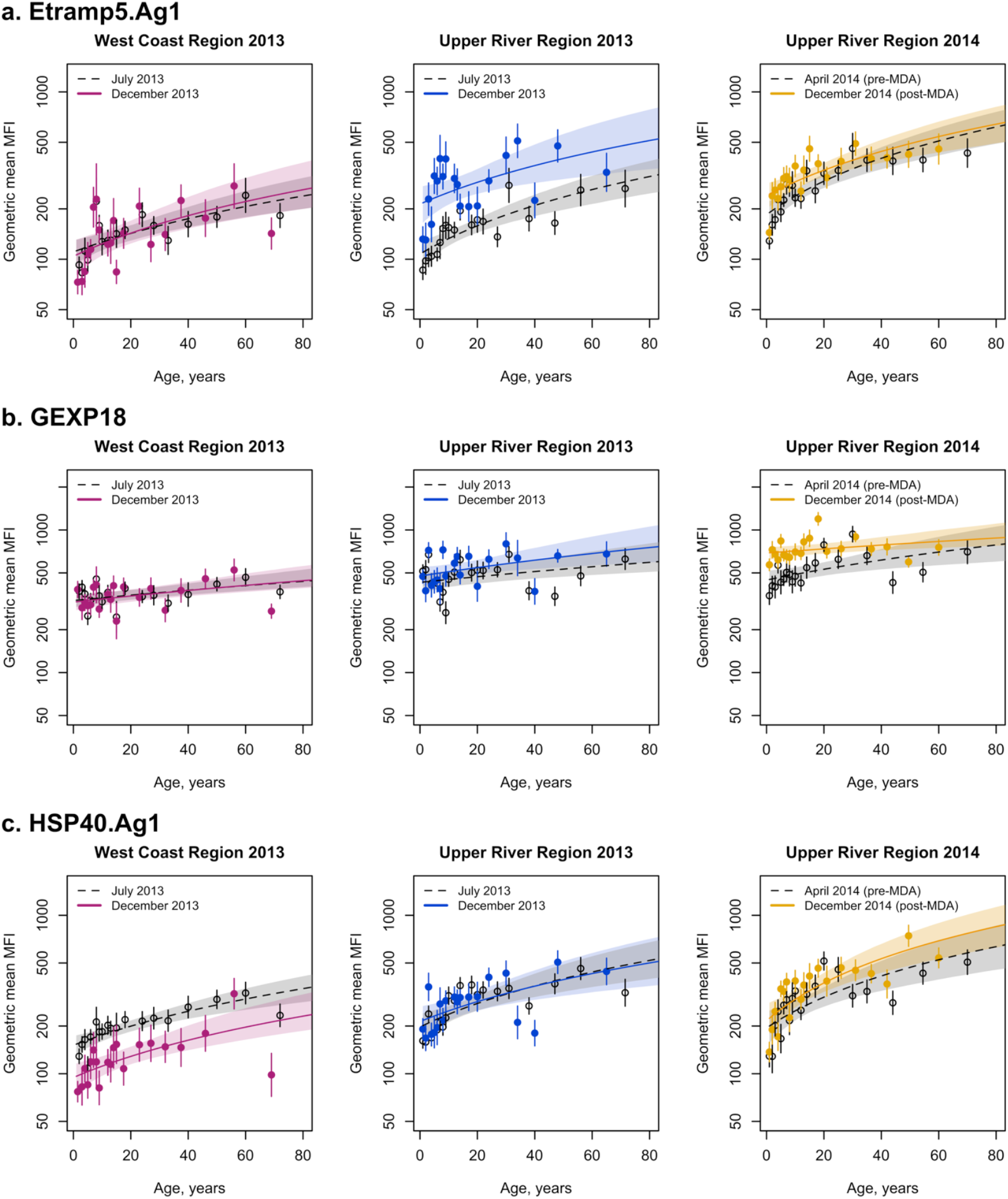
Age-dependent antibody acquisition for Etramp5.Ag1, GEXP18 and HSP40.Ag1 by geographical region and transmission season. Mean and 95% confidence intervals of geometric mean antibody levels for each age group are shown as circles and vertical lines, respectively. Median fit of the antibody acquisition model is shown as a solid line for the end of the transmission season (December 2013 and 2014) and dotted line for the dry season (April 2014) or start of the transmission season (July 2013). Shaded regions are the 95% credible intervals of the model fit.

At the start of the 2013 malaria transmission season (July), *Pf*MSP1^19^ SCR in URR was twice as high (SCR 0·0130, 95%CI 0·0088 – 0·0194) than in WCR (SCR 0·0067, 95%CI 0·0037 – 0·0123). After peak transmission season, this difference was more pronounced, with *Pf*MSP1^19^ SCR 0·0214 (95%CI 0·0087 – 0·0530) in WCR compared to SCR 0·0535 (95%CI 0·0344 – 0·0831) in URR. *Pf*AMA1 SCR in the URR was consistently higher than the WCR throughout the transmission season, with SCR 0·0159 (95%CI 0·0080 – 0·0239) and 0·0614 (95%CI 0·0509 – 0·0740) for WCR and URR, respectively, in July, and SCR 0·0166 (95%CI 0·0124 – 0·0224) and 0·0915 (95%CI 0·0738 – 0·1135) in WCR and URR, respectively, in December. Increases in sero-prevalence in older age groups are not pronounced for markers associated with short-lived antibody responses, which is an underlying assumption when using sero-catalytic models. Therefore, antibody acquisition models are used to assess age-adjusted changes in antibody responses based on continuous data for markers of short-lived antibody responses.

### Age-adjusted antibody acquisition by geographical region and transmission season

For antigens associated with long-lived antibody responses (*Pf*MSP1^19^ and *Pf*AMA1), differences in age-adjusted antibody acquisition were observed between WCR and URR at the end of the 2013 transmission season (December) (Figure 5, Supplementary Tables 8-9). For *Pf*MSP1^19,^ differences in geometric mean antibody levels in children aged 1-15 years in URR were higher at the end of the transmission season compared to the start of the season (p=0·03). Additionally, higher antibody levels to *Pf*AMA1 in 1-15 year olds were observed in URR compared to WCR at the end of the transmission season (p<0·001). No significant differences in antibody levels for *Pf*AMA1 were observed between the start and end of transmission season in URR.

**Figure 5.**
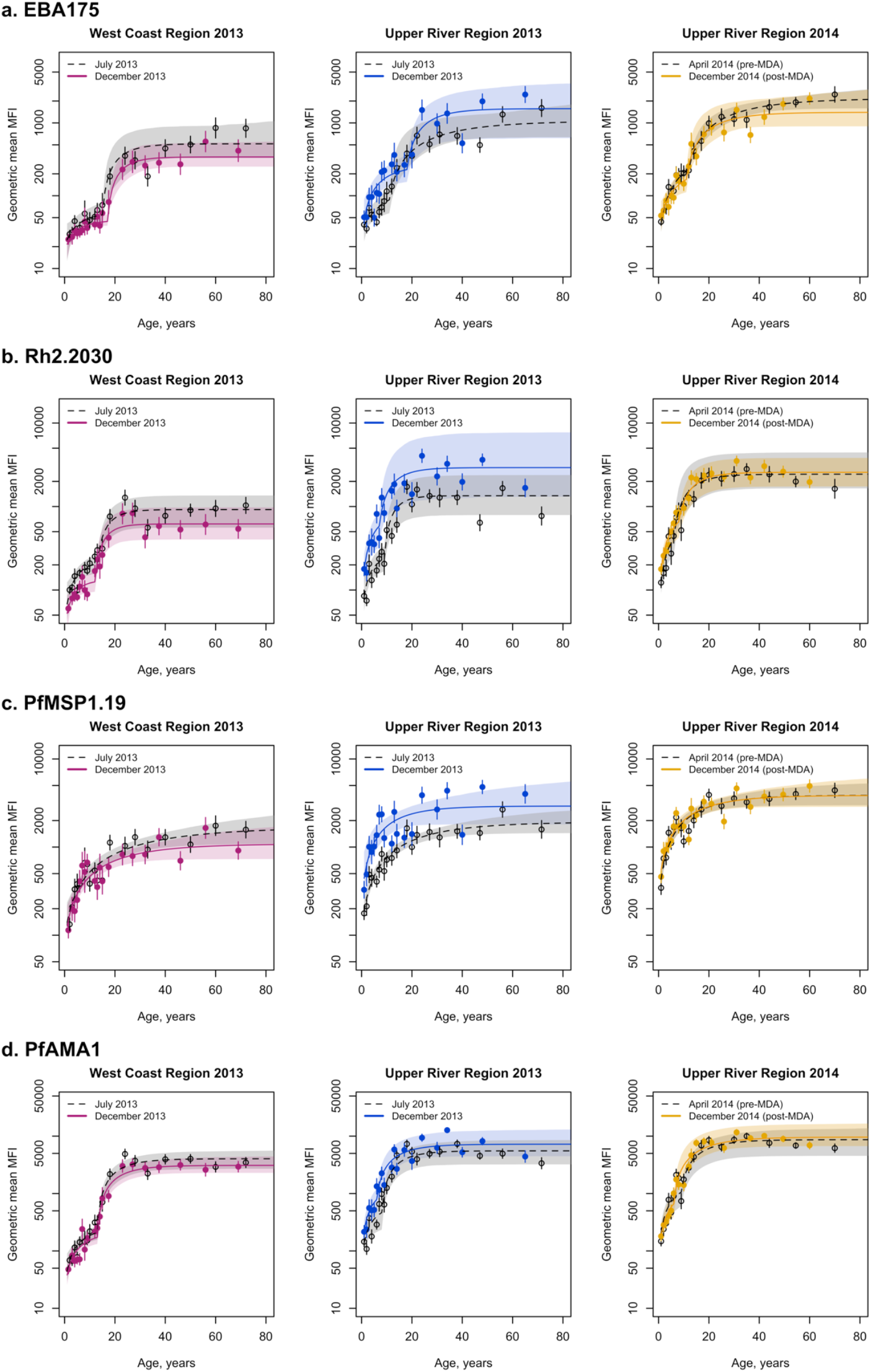
Age-dependent antibody acquisition for EBA175, Rh2.2030, *Pf*MSP1.19 and *Pf*AMA1 by geographical region and transmission season. Mean and 95% confidence intervals of geometric mean antibody levels for each age group are shown as circles and vertical lines, respectively. Median fit of the antibody acquisition model is shown as a solid line for the end of the transmission season (December 2013 and 2014) and dotted line for the dry season (April 2014) or start of the transmission season (July 2013). Shaded regions are the 95% credible intervals of the model fit.

In contrast to *Pf*MSP1^19^ and *Pf*AMA1, all serological markers associated with shorter-lived antibody responses (Etramp5.Ag1, GEXP18, HSP.Ag1, EBA175, and Rh2.2030) showed higher antibody levels in 1-15 year olds in URR compared to WCR at the end of the transmission season (Figure 4, Supplementary Tables 3-7). In URR, antibody responses to Etramp5.Ag1 increased between the start and end of the transmission season amongst individuals 1-15 years (p=0·001). However, antibody levels in 1-15 year olds for HSP40.Ag1 decreased between the start and end of the transmission season in WCR (p=0·004). At the start of the transmission season, antibody levels to GEXP18 were higher in URR compared to WCR (p=0·03). There were no within-region differences in antibody responses to any of the antigens tested between the 2013 transmission season and the following dry season (April 2014).

### Association between pre-MDA antibody levels and post-MDA malaria infection

Decreases in under-15 sero-prevalence between pre- and post-MDA transmission seasons (December 2013 vs. December 2014) were only observed for GEXP18 (diff-3·4% 95%CI −9·2, 2·4) and HSP40.Ag1 (diff-2·1% 95%CI −7·0, 2·9), but differences were not statistically significant. After adjusting for age, LLIN use in the last 24 hours, and MDA compliance, individuals in the upper 50^th^ percentile of above-average antibody levels to Etramp5.Ag1 for their age group in the dry season (April 2014, before the implementation of MDA) had two-fold higher odds of clinical malaria (aOR 1·91 95%CI 1·05 − 3·52, p=0·04) in the 2014 transmission season (October – December) after MDA (Figure 6A, Supplementary Table 10). Dry season antibody levels to Rh2.2030 were also moderately associated with increased odds of clinical malaria (OR 1·75 95%CI 0·93 - 3·30, p=0·09) after MDA (Figure 6A, Supplementary Table 14), but statistical evidence was not strong. Similarly, there was a borderline association between above average antibody levels to *Pf*AMA1 in the dry season and increased odds of clinical malaria (OR 1·70 95%CI 0·97 - 2·98, p=0·07), but statistical association was weak after adjusting for age, LLIN use and MDA compliancy (Figures 6A, Supplementary Table 16). Antibody responses to all other antigens investigated were not associated with increased odds of infection post-MDA (Supplementary Tables 11-15).

**Figure 6.**
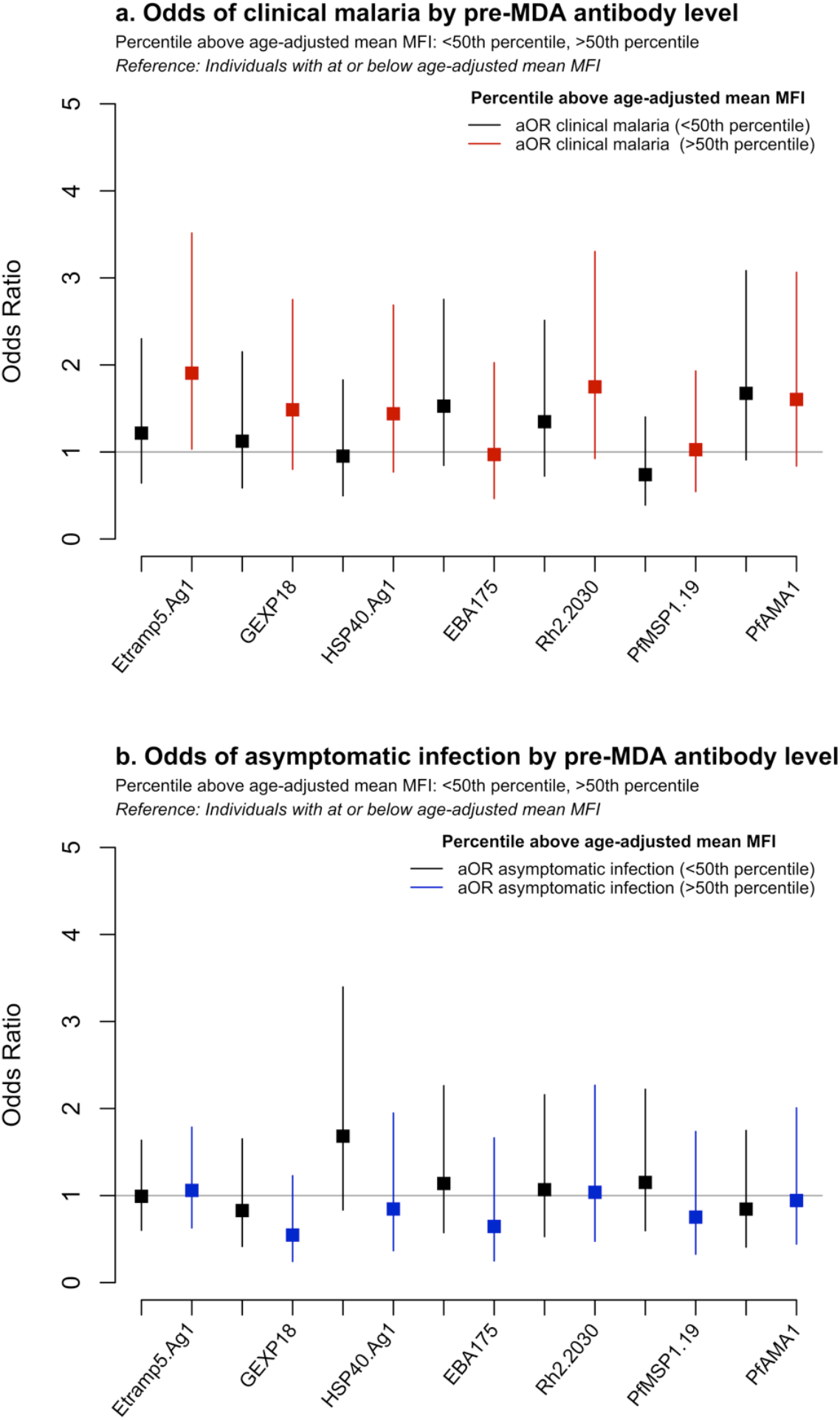
Association of pre-MDA dry season (April 2014) antibody levels and odds of (a) clinical malaria or (b) asymptomatic infection, during transmission season post-MDA (December 2014). Odds ratios are adjusted for age, LLIN use and MDA compliancy. Clinical malaria is defined as clinical or RDT confirmed case, asymptomatic infection is defined as PCR-positive and clinically or RDT-negative.

## Discussion

In this study, spatio-temporal variations in serological responses were observed for several *Pf* antigens, likely reflecting changes in malaria incidence as well as residual antibodies from prior infection. In moderate to high transmission settings, higher antibody levels in the dry season due to prior exposure are common. However, the rate at which protective antibodies develop varies by setting. Immunity to clinical malaria has been observed in children as young as 5 years^32,33^, while in other transmission ranges, immunity may only develop after 10 to 15 years of exposure.^34,35^ Children can be a useful sentinel population for monitoring transmission, but the optimal age range for surveillance depends on the historical patterns of transmission governing population antibody responses. Most indicators and survey designs are associated with some bias affecting the accuracy of estimates. School surveys in South East Asia, where the distribution of malaria may be skewed towards older age groups, may better reflect sero-incidence due to less childhood exposure compared to sub-Saharan Africa. Serological surveillance protocols can therefore be combined with other routine metrics to overcome potential sampling biases.

Differences in sero-conversion rates to *Pf*MSP1^19^ and *Pf*AMA1 were observed between geographical regions but not between transmission seasons. Long-lived antibody responses typically reflect past exposure rather than incidence during a single transmission season. However, these markers have been shown to be sensitive to changes in transmission over longer periods of time in The Gambia.^36^ Antibody acquisition models can detect more subtle differences in the magnitude of antibody levels. Five sero-incidence markers evaluated in this study - Etramp5.Ag1, GEXP18, HSP40.Ag1, EBA175, and Rh2.2030 - detected geographical differences in antibody acquisition. Additionally, Etramp5.Ag1 and HSP40.Ag1 detected seasonal differences in high and low transmission settings. These findings align with previous microarray studies, where Etramp5.Ag1, GEXP18, HSP40 were amongst the top 10 serological markers out of 856 *Pf* antigens screened found to be most associated with recent episodes of clinical malaria in Uganda and Mali^21^.

Between December 2013 and April 2014, antibody levels did not decline significantly, indicating that antibody decay probably occurs over a period longer than four months. This may be due to persistent asymptomatic parasitaemia or infections occurring late in the transmission season. In The Gambia, antibody decay in children has been observed for several antigens associated with protective immunity (*Pf*AMA1, *Pf*MSP2, *Pf*MSP1, EBA175), with antibody half-life estimated between 10 to 50 days^37,38^ and long-lived antibody secreting cells persisting for up to four years^38^. However, antibody half-lives vary between settings, ranging from 10-20 days in Kenya^39^ and Ghana^38^, 98 to 120 days in Nigeria for EBA175 and Rh4.2^40^, and up to 225 days in Cambodia^41^ and 7 years in Thailand^42^ for *Pf*MSP1^19^. However, these studies focus on a limited number of antigens. Estimates of antibody longevity for a larger range of antigens can help inform optimal sampling strategies for future serological surveillance using multiplex assay platforms. Studies in Mali have quantified antibody responses to over 2,000 *Pf* antigens on protein microarray, observing rapid declines in antibody levels within six months.^43^

Above average antibody levels in the dry season before MDA was associated with increased odds of clinical malaria following MDA. Associations were moderate for Rh2.2030 and *Pf*AMA1, but individuals with higher than average antibody levels against Etramp5.Ag1 had a two-fold higher odds of clinical malaria post-MDA. It is possible that serology can identify individuals who are infected during the dry season or experience a high frequency of exposure, leading to increased risk of clinical illness, despite remaining below the detection limit of other diagnostics. Similar associations were observed using parasitological endpoints, where individuals with *Pf* infection as measured by PCR in the dry season had increased odds of infection post MDA.^22^ This study was not designed to assess the effectiveness of MDA using serological endpoints, and sample size may not be powered to draw strong conclusions. Analysis of a larger set of antigens or grouping antibody data based on antigen expression or function during the parasite life cycle may help delineate the wide range of responses with respect to malaria exposure, incidence, or immunity. Studies measuring persistent asymptomatic parasitaemia throughout the dry and wet seasons could further elucidate the dynamics between parasite density, immunity, and serological responses.

This study demonstrates that a diverse panel of serological markers can be used to monitor malaria exposure (observed through long-lived antibody responses) and incidence (through shorter-lived antibody responses). Microarray studies evaluating the combined use of multiple serological markers suggest that panels of at least five antigens confer greater diagnostic accuracy compared to a single marker, while panels of more than 20 have only incremental improvements in accuracy^21,44^. Between-antigen variation in antibody detection will also exist naturally; differences in the immunogenicity of recombinant antigen constructs could be due to sequence selection or expression systems, or the choice of antigen isotypes or IgG subclasses. Multi-marker diagnostic panels may be better able to capture the breadth of antibody responses in the population. This lends strong support for the combined use of *Pf* antigens identified in this study in future diagnostic platforms. Evaluating their accuracy in future intervention trials and surveillance studies, particularly cluster randomised trials, can help establish standardised serological protocols usable across a variety of epidemiological settings.

## Data Availability

The datasets used and/or analysed during the current study are available from the corresponding author on reasonable request.

## List of abbreviations

aOR: Adjusted odds ratio
AUC: Area Under the Antibody Acquisition Curve
CI: Confidence interval
DBS: Dried blood spot
DHA-PQ: Dihydroartemisnin-piperaquine
EBA175: Erythrocyte binding antigen 175 RIII-V
Etramp5.Ag1: Early transcribed membrane protein 5
GEE: Generalised estimating equations
GEXP18: Gametocyte export protein 18 (GEXP18)
HSP40.Ag1: Heat shock protein 40
IgG: Immunoglobulin
LLIN: Long-lasting insecticide treated net
MDA: Mass drug administration (MDA)
MFI: Median fluorescence intensity
PCD: Passive case detection
PCR: Polymerase chain reaction
*Pf, P*.*falciparum*: *Plasmodium falciparum*
*Pf*AMA1: *P*.*falciparum* apical membrane antigen 1
*Pf*MSP1^19^: *P*.*falciparum* merozoite surface antigen 1 19-kDa carboxy-terminal region
RDT: Rapid diagnostic test
Rh2.2030: Reticulocyte binding protein homologue 2
SCR: Sero-conversion rate
URR: Upper River Region, The Gambia
WCR: West Coast Region, The Gambia

## Declaration of interests

### Ethics approval and consent to participate

This study was approved by the Gambia Government/MRC Joint Ethics Committee (SCC1318). Verbal consent was first obtained at village sensitisation meetings, followed by individual written informed consent for all participants. Parents/guardians provided written consent for children less than 17 years, and assent was obtained from children between 12 and 17 years.

### Author contributions

LW designed and coordinated the study, supervised and performed serological lab work, analysed the data, and wrote the manuscript. JM and MA supervised field data collection and laboratory work. SC and MB conducted the serological assays. KKAT and JB produced the antigens. KKAT and TH developed the serological assay. JM, MA, UD, IK, and CD advised on study design, interpretation of findings, and reviewed the manuscript. All authors read and approved the final manuscript.

